# Prevalence and Incidence of Antibodies Against Sars-Cov-2 Among Primary Healthcare Providers in Belgium During One Year of the Covid-19 Epidemic: Prospective Cohort Study Protocol

**DOI:** 10.1101/2021.06.18.21259139

**Authors:** Niels Adriaenssens, Beatrice Scholtes, Robin Bruyndonckx, Jan Y Verbakel, An De Sutter, Stefan Heytens, Ann Van den Bruel, Isabelle Desombere, Pierre Van Damme, Herman Goossens, Laëtitia Buret, Els Duysburgh, Samuel Coenen

## Abstract

**Introduction:** National severe acute respiratory syndrome coronavirus 2 (SARS-CoV-2) seroprevalence data provides essential information about population exposure to the virus and helps predict the future course of the epidemic. Early cohort studies have suggested declines in levels of antibodies in individuals, associated with, for example, illness severity, age and co-morbidities. This protocol focuses on the seroprevalence among primary health care providers (PHCPs) in Belgium. They manage the vast majority of COVID-19 patients in addition to other patients and therefore play an essential role in the efficient organisation of health care. Currently, evidence is lacking on 1. how many PHCPs get infected with SARS-CoV-2 in Belgium, 2. the rate at which this happens, 3. their clinical spectrum, 4. their risk factors, 5. the effectiveness of the measures to prevent infection and 6. the accuracy of the serology-based point-of-care test in a primary care setting.

**Methods and analysis:** This study will be set up as a prospective cohort study. General practitioners (GPs) and other PHCPs (working in a GP practice) will be recruited via professional networks and professional media outlets to register online to participate. Registered GPs and other PHCPs will be asked at each testing point (n=9) to perform a capillary blood sample antibody point-of-care test (OrientGene®) and complete an online questionnaire. The primary outcomes are the prevalence and incidence of antibodies against SARS-CoV-2 in PHCPs during a 12-month follow-up period. Secondary outcomes include the longevity of antibodies against SARS-CoV-2.

**Ethics and dissemination:** Ethical approval has been granted by the Ethics Committee of the University Hospital Antwerp/University of Antwerp (Belgian registration number: 3002020000237). Alongside journal publications, dissemination activities include the publication of monthly reports to be shared with the participants and the general population through the publicly available website of the Belgian health authorities (Sciensano).

**Registration:** Trial registration number: NCT04779424

**Article summary:** *Strengths and limitations of this study:*

- This large cohort study will provide regular, timely and precise data at national level on prevalence and incidence of antibodies against SARS-CoV-2 among primary health care providers (PHCPs) managing the vast majority of COVID-19 and other patients and therefore essential to organise health care efficiently.
- This study will familiarise PHCPs with the use of serology-based point-of-care tests (POCTs) and validate the POCT in a primary care setting.
- Missing data points and the use of a convenience sample could limit the validity of the study results.

## Introduction

As of 16th May 2021, severe acute respiratory syndrome coronavirus 2 (SARS-CoV-2) has infected over 162 million people worldwide (over 1 030 000 in Belgium) and caused over 3.3 million deaths from coronavirus disease (COVID-19) worldwide (over 24 000 in Belgium).^1^ COVID-19 is a lethal respiratory tract infection (RTI), but infection with SARS-CoV-2 can also be mild and even asymptomatic.

SARS-CoV-2 seroprevalence estimates provide essential information about population exposure to infection and help predict the future course of the epidemic.^2 3^ When setting up this study seroprevalence studies in Iceland^4^ and Spain^5^ showed different levels of population antibody positivity, lasting up to 4 months in Iceland. In addition, cohort studies have suggested substantial waning of antibody levels in individuals, associated with for example illness severity, age and co-morbidities.^6-8^ Meanwhile, other seroprevalence studies showed antibody positivity lasting up to 9 months.^9 10^ For Belgium, Sciensano (the Belgian national scientific institute, www.sciensano.be) performs national seroprevalence studies of SARS-CoV-2 antibodies in several relevant populations including schools,^11^ hospital personnel^12^ and nursing homes^13^.

This protocol focuses on the seroprevalence among primary care health care providers (PHCPs). They manage the vast majority of COVID-19 and other patients and therefore play an essential role in the efficient organisation of healthcare.^14 15^ Among the PHCPs, general practitioners (GPs) in particular, act as gatekeepers to the next levels of care. Therefore, preserving the capacity of GPs, together with that of their co-workers, throughout the COVID-19 epidemic is essential.^16^ In Belgium, this is particularly concerning given the GP workforce consists of mainly older adults and is therefore at higher risk for COVID-19-related morbidity and mortality.^17^ In Italy GPs represented up to 38% of the physicians who died from COVID-19 early on in the epidemic.^18^

However, current evidence is lacking on 1. how many PHCPs are infected by SARS-CoV-2 or have COVID-19 in Belgium, 2. the rate at which this occurs, 3. their clinical spectrum, 4. their risk factors, 5. the effectiveness of the measures to prevent this from happening and 6. the accuracy of the immunological serology-based point-of-care test (POCT) used by PHCPs.

During the COVID-19 crisis POCTs have been developed to identify the presence of antibodies for SARS-CoV-2. Compared to laboratory tests, a valid easy-to-use POCT will speed up the availability of the test results for both the participants and the national health authorities. Furthermore, by using POCTs in this study, PHCPs will have the opportunity to become more familiar with this type of technology.

Sciensano has validated five POCTs using finger prick blood, identifying one test with appropriate sensitivity (92.9%) and specificity (96.3%) for use in seroprevalence studies.^19^ We use this Orientgene® POCT for the present study.

If (Belgian) primary care cannot be delivered safely, the COVID-19 epidemic will disrupt public health by failing to deliver non-COVID-19 related healthcare and to (continue to) keep off the pressure from the next levels of care during the current epidemic. Therefore, we need to monitor their health and the effectiveness of, and the need for, infection prevention and control measures during epidemics. In addition, the follow-up of a cohort of PHCPs will help us to understand the duration and nature of antibodies generated in response to SARS-CoV-2 infection as well as those generated in response to vaccination.^20^ Whether and for how long antibody response protects those infected with SARS-CoV-2 from future infections or illness will determine the value of serological tests.^21^

### Primary objectives

1. Assess the prevalence of antibodies against SARS-CoV-2 among PHCPs (PHCPs = GPs and other PHCPs in their practice) in Belgium at timepoint 1 and at different timepoints during a 12-month follow-up period.
2. Assess the monthly and annual incidence of antibodies against SARS-CoV-2 among PHCPs in Belgium during a 12-month follow-up period

### Secondary objectives

1. Assess the longevity of the serological antibody response among seropositive PHCPs.
2. Assess the proportion of asymptomatic cases among (new) cases (that develop during follow-up).
3. Assess the determinants (risk and predictive factors) of SARS-CoV-2 infection in PHCPs.
4. Validate the serology-based POCT in a primary care setting (Phase 3 validation).
5. Familiarise PHCPs with the use of serology-based point-of-care tests.

Once vaccination of PCHPs starts, this study will take into account vaccination rates when reporting the seroprevalence and be able to assess waning of antibodies after vaccination.

## Methods and analysis

The aim of this study is to broaden the knowledge on SARS-CoV-2 infection in Belgian primary care and to contribute to scientific research, health service and policy management supporting the fight against this epidemic.

### STUDY POPULATION

#### Inclusion Criteria

– Any GP working in Belgium (including those in professional training) currently working in primary care and any PHCP from the same GP practice who physically manages (examines, tests, treats) patients,
– Participants must be able to comply with the study protocol and provide informed consent to participate in the study.

#### Exclusion Criteria

– Staff hired on a temporary (interim) basis will be excluded as follow-up over time will be compromised.
– Administrative staff or technical staff without any prolonged (longer than 15 minutes) face to face contact with patients are not eligible.
– PHCPs who were not professionally active during the inclusion period will not be eligible

### STUDY DESIGN

This study will be set up as a prospective cohort study.

#### Recruitment

PHCPs will be recruited prior to the first and second testing points (registration will be possible between 15 November 2020 and 15 January 2021). PHCPs working in clinical practice in Belgium will be invited to register online for participation in this national epidemiological study and will be asked to invite the other PHCPs in their practice to do the same. We will emphasize that PHCPs that have already been diagnosed with COVID-19 are also eligible. Information about the study will be disseminated to GPs and PHCPs via professional organisations (Domus Medica and College de Médecine Générale), university networks across the country and through professional media channels. The convenience sample of participants will be checked to ensure that it is representative in terms of geographic and demographic qualities.^22^

#### Data Collection

Upon inclusion in the study, participants will be assigned a unique study code by the researchers, who will manage the key between these codes and the identification data. They will receive testing material at their place of work through regular mail. At the first testing time-point (T1) they will receive an invitation by email (including a personalised link to an online questionnaire in French and Dutch) inviting them to:

1. Auto-collect a capillary blood sample and analyse it using the OrientGene® POCT.
2. Complete a baseline questionnaire through a secured online platform hosted by Sciensano (Limesurvey).

The baseline questionnaire at the first testing point will ask for their informed consent and will ask for information about;

– The result of the POCT,
– basic socio-demographic data, (age, gender, composition of household – e.g. presence of school-aged children in the house)
– professional data, (practice patient size)
– health status, (pre-existing health conditions, regular medication use, presence of symptoms since the start of the epidemic, previous positive test results for COVID-19)
– Professional exposure, (contact with confirmed cases, use of infection prevention and control measures and
– the availability of personal protective equipment (practice organisational aspects, delayed care for non-urgent conditions) (see supplementary materials).

A follow-up questionnaire will be sent for each of the subsequent testing timepoints. In addition to the POCT result, it will collect information on:

– the health status, including the presence of symptoms,
– vaccination status (date of vaccination, type of vaccine, number of doses, presence of side-effects)
– professional exposure, (contact with confirmed cases, use of infection prevention and control measures (see supplementary materials).

#### Phase 3 validation of the POCT

To validate the POCT, a sub-sample of participants will be asked to provide a serum sample. This sub-sample will be made up of all those participants that were seropositive for SARS-Cov-2 on the POCT at T1 and a random sample of participants that were seronegative at T1.

The participants will be sent material to collect the blood sample (Becton Dickinson Vacutainer® SSTTM ii Advance; ref 368879) along with postal materials (in accordance with the UN 3373 packaging norms) and instructions on how to send it to the laboratory of clinical biology of the University Hospital of Antwerp (UZA) (a reference laboratory chosen by Sciensano). Participants will be asked to send their blood sample the same day it is taken, and analysis will be undertaken within 24 hours of reception. Analysis will be done with a reference standard using the following testing algorithm: serum samples will be tested first on the ELECSYS Anti-SARS-CoV-2 S assay (Roche, Basel, Switzerland), if the cut-off index (COI) is between 0.6-3.0 the sample will be tested on the ATELLICA IM SARS-CoV-2 assay (Siemens, Munich, Germany), and if discordant results it will be tested on the LIAISON SARS-CoV-2 S1/S2 IgG assay (DiaSorin, Saluggia, Italy), using a two out of three ‘reference standard’. The analytical and clinical performance of these three commercially available, fully automated SARS-CoV-2 antibody assays was investigated at University hospital of Antwerp (UZA) and the relevance of this testing algorithm explained and illustrated (personal communication Bart Peeters). Analytical performance of all three assays was acceptable and comparable with results found in other studies.^23-26^

Participants of this sub-sample will receive a 25€ voucher by way of compensation for the time and effort invested. The results of the serum sample will be communicated to participants via regular mail.

#### Follow-up

The study will last 12 months. Epidemiological data collected through the online questionnaires and self-sampling using the POCT will occur monthly for six months with one sample collection at nine and the final one at 12 months (Table 1). This corresponds to a total of nine testing timepoints. This number will however depend on the evolution of the epidemic. At each testing time-point, participants will be asked to perform the POCT within a timeframe of maximum 5 days. For the sub-sample providing a serum sample, participants will be asked to take the serum sample at the same time (just prior) to performing the POCT.

**Table 1.**
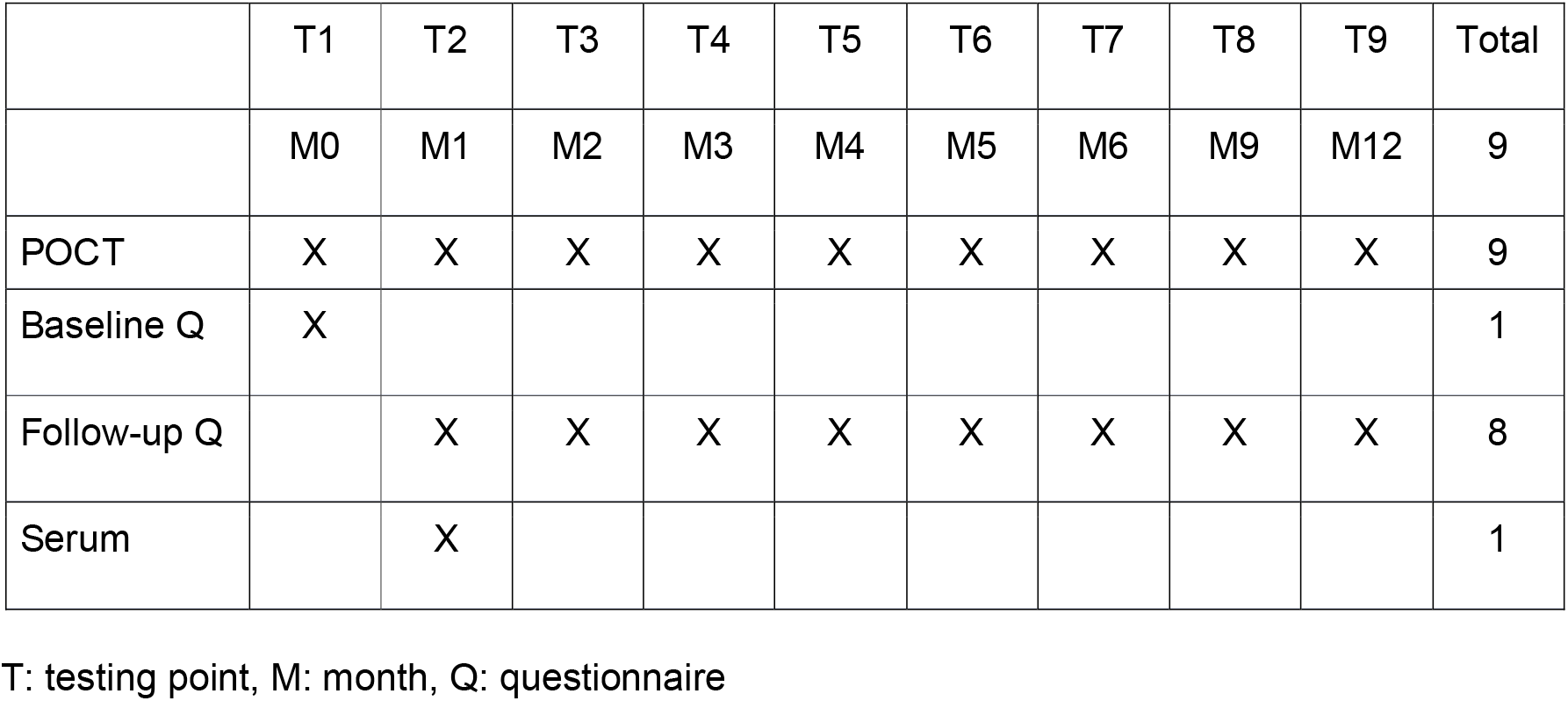
Timing of data collection.

|  | T1 | T2 | T3 | T4 | T5 | T6 | T7 | T8 | T9 | Total |
| --- | --- | --- | --- | --- | --- | --- | --- | --- | --- | --- |
|  | M0 | M1 | M2 | M3 | M4 | M5 | M6 | M9 | M12 | 9 |
| POCT | X | X | X | X | X | X | X | X | X | 9 |
| Baseline Q | X |  |  |  |  |  |  |  |  | 1 |
| Follow-up Q |  | X | X | X | X | X | X | X | X | 8 |
| Serum |  | X |  |  |  |  |  |  |  | 1 |

T: testing point, M: month, Q: questionnaire

The result of the POCT will be entered as a variable in the online questionnaire.

Data analysis will be performed and reported after each relevant testing period and at the end of the study. All pseudonymised data collected will safely be stored by Sciensano for 10 years after completion of the study.

### SAMPLE SIZE

This study aims to include 5000 PHCPs with a 4 GPs to 1 other PHCP ratio considering the following sample size considerations regarding the different objectives of the proposed study.

To estimate a prevalence ranging from 5% to 10%, the current estimates for SARS-CoV-2 seroprevalence in the general population and hospital care providers, with a precision ranging from 2% to 1% and a 95% confidence level, a sample size ranging from 504 to 3554 PHCPs is required (Binomial ‘exact’ calculation), respectively. Since PHCPs will be clustered in their practices, we must correct the sample size. For an average of 2.5 PHCPs per practice (m) and an intraclustercorrelation of 0.2 (rho) the design effect (=1+(m-1)*rho) is 1.3. The corrected sample size ranges from 655 to 4620 PHCPs. Higher seroprevalence and non-response, both of which are to be expected, will reduce the precision of the estimates as will stratification by region or province. For example, with a sample size of 4620 PHCPs distributed equally over eleven strata, which corresponds to the number of provinces in Belgium (n=10) plus Brussels, the precision will range between 2.5% and 3.5% for a prevalence ranging from 5% to 10%, respectively.

Since multivariate prediction research for each determinant studied requires at least 10 subjects in the smallest category of the outcome variable to allow proper statistical modelling,^27 28^ a model including 25 determinants would require 250 seropositive participants, which corresponds to a 5% seroprevalence in 5000 or a 10% seroprevalence in 2500 PHCPs, not taking into account interaction terms in the model. The number of determinants that can be assessed in multivariable analysis to predict new cases will depend on the incidence. For example, to be able to assess 10 determinants would require 100 new cases or 3% new cases in 3600 PHCPs or 4700 PHCPs considering a design effect of 1.3. A lower incidence or lower sample size would further limit the number of determinants that can be modelled. Using more recently described methods to calculate the sample size required for developing a clinical prediction model would also require a sample size of substantially more than 2000 participants (n=2283, with 228 events and 9.1 events per predictor) to meet the 4 criteria described by Riley RD et al. in case of a Mean Average Prediction Error (MAPE) of 0.025.^29^

To estimate an incidence of 3% with a precision of 1% and a 95% confidence level, a sample size of 1212 PHCPs is required or 1576 PHCPs considering a design effect of 1.3 (4160 PHCPs to estimate an incidence of 2% with a precision of 0.5% and considering clustering). To be able to validate the POCT’s accuracy in the primary care setting, i.e. estimate the POCT’s sensitivity (92.9%) with a lower limit of its 95%CI of 90% and its specificity (96.3%) with a lower limit of its 95%CI of 95%, a sample of 301 PHCPs seropositive on the reference standard (for sensitivity) and 810 PHCPs seronegative on the reference standard (for specificity) is required, which corresponds to for example 6% seroprevalence in 5022 PHCPs. To reduce the burden on the participants and the costs of the study all those with a positive POCT and only a (random) sample of 900 PHCPs with a negative POCT will be assessed with the reference standard, and inverse probability weighting will be applied to correct for missing reference standard data by design.^30-32^

A sample size of 5000 would also allow us to estimate the longevity of the antibody response among the PHCPs seropositive on the POCT. For example, starting from 300 PHCPs seropositive based on the POCT, a decrease of 10% in seroprevalence can be estimated with a precision of 4% and a 95% confidence level. Smaller decreases in seroprevalence and/or estimating with lower precision would require less than 5000 PHCPs to identify sufficient PHCPs seropositive on the POCT. Clustering will most likely not be an issue here since the waning of antibodies will most likely not be correlated among PHCPs working in the same practice.

### DATA PROTECTION

As described above, epidemiological and serological data will be linked via a unique identifier code assigned to each participant. The same unique identifier code will be entered in each questionnaire, enabling the link for data analysis. This code will stay the same throughout the study. The key between the codes and the identification data of the participants will be kept in a secure and protected way by the principal investigators and the researchers, and destroyed upon completion of the study. The personal data processing activities for the proposed research project will be submitted to the UAntwerpen Data Protection Office to review its completeness and compliance with the General Data Protection Regulation (GDPR) and to ask for formal approval. To control digital access only by authorized people on all devices (desktops, laptops, external drives, …) at all locations (work, home, and travel), complex passwords are used, up-to-date anti-virus and firewall protection is run. Using the ICT services of UAntwerp, ULiège and Sciensano assures that the data will be backed up on a regular basis. The research team ensures that their personal computer system is always up-to-date, and does not switch off the automatic installation of updates.

### DATA ANALYSIS

Data analysis will be done jointly by the principal investigators, researchers and team involved in this study with the University of Antwerp team taking the lead. Questionnaire responses will be coded. Data will be cleaned and validated; incomplete questionnaires will be manually checked to see if they can be included. Analysis will be mainly descriptive and done on R version 3.6.3 or equivalent.

Among others, the following indicators will be calculated, considering clustering of PHCPs in the same practice whenever appropriate:

1. Seroprevalence of SARS-CoV-2: number of participants in whom presence of specific SARS-CoV-2 IgG is detected by the POCT / Total number of participants tested with the POCT
2. Prevalence of reported COVID-19 cases: number of participants who self-report at baseline that SARS-CoV-2 infection (symptomatic and asymptomatic) was detected / Total number of participants responding to the baseline questionnaire
3. SARS-CoV-2 seroconversion rate: number of participants in whom presence of specific SARS-CoV-2 IgM and/or IgG is detected by POCT at follow-up / Total number of participants followed-up not sero-converted before (based on prior POCT results), monthly during 12 months of follow-up.
4. Incidence of reported COVID-19: number of participants who self-report new SARS-CoV-2 infections (symptomatic and asymptomatic) at follow-up / Total number of participants not yet infected before (based on prior self-reporting and POCT results) and responding to the follow-up questionnaire, monthly during 12 months of follow-up.
5. SARS-CoV-2 antibodies longevity: number of participants in whom presence of specific SARS-CoV-2 IgG is no longer detected by POCT at follow-up / Total number of participants followed-up sero-converted before (based on prior POCT results), monthly during 12 months of follow-up.

To assess determinants of SARS-CoV-2 seroprevalence and seroconversion in PHCPs, among which the availability and use of different preventive measures against SARS-CoV-2 infection, univariable and multivariable regression analysis, considering the clustering of participants at their practices, will be performed, e.g. generalised estimating equations.^33^ Model calibration will be assessed using calibration plots and the Hosmer-Lemeshow goodness-of-fit test.^34^ Its discrimination will be estimated with the area under the receiving operator characteristic (ROC) curve.

### Data Analysis Phase 3 validation POCT

To validate the POCT in a primary care setting, we will estimate the following test characteristics:

1. SARS-CoV-2 POCT sensitivity: number of participants testing positive on the SARS-CoV-2 POCT / Total number of participants testing positive on the reference standard.
2. SARS-CoV-2 POCT specificity: number of participants testing negative on the SARS-CoV-2 POCT / Total number of participants testing negative on the reference standard.

These estimates will be corrected for missing reference standard data by inverse probability weighting to infer what the reference standard results might have been had the entire study sample been verified.^30-32^ To show which participants are missing a reference standard result a flow chart will be provided (Figure 1).

**Figure 1.**
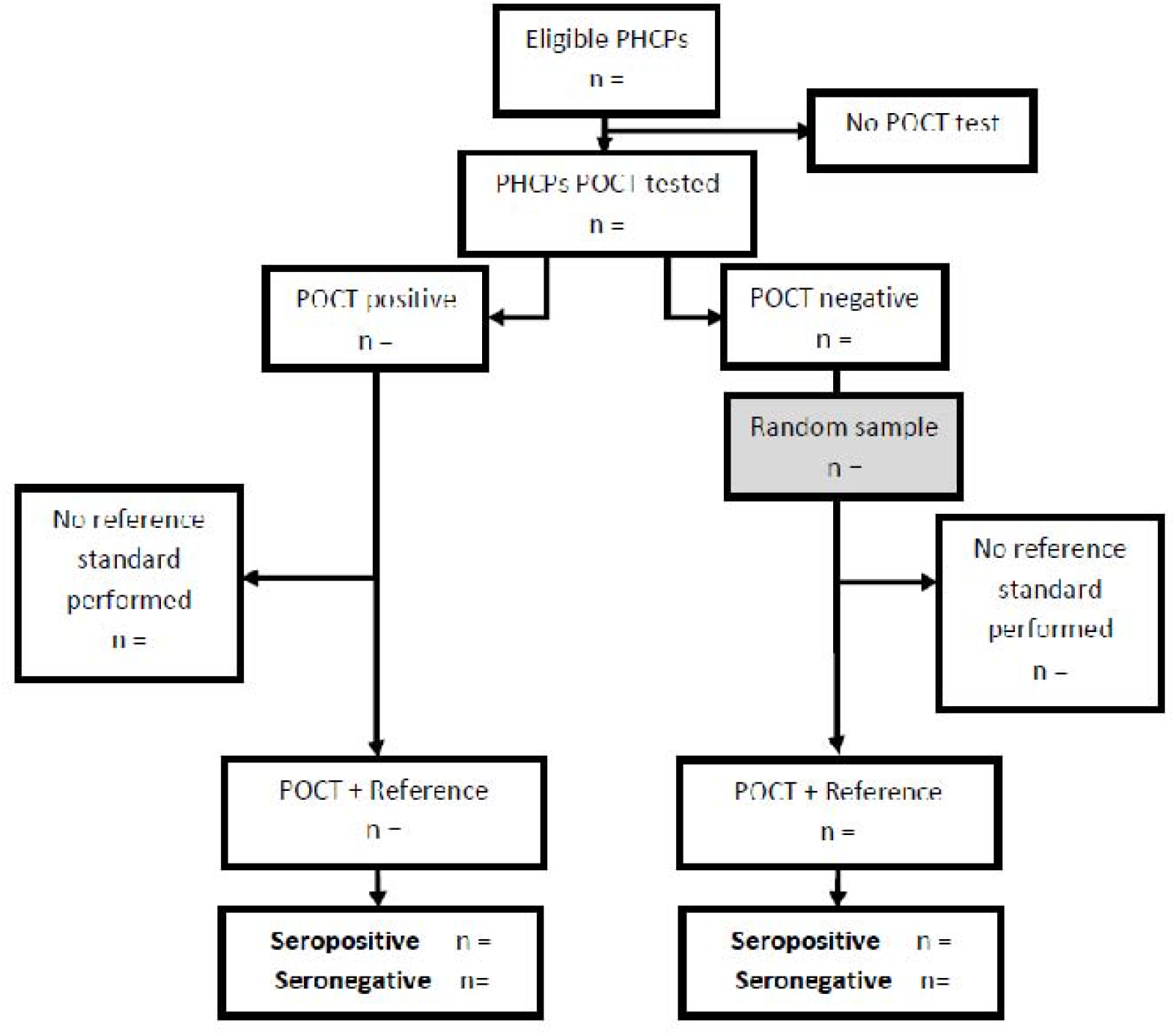
Participant flow

### Vaccination

The start of the vaccination of PHCPs during the study follow-up provided the opportunity to monitor its progress (at regional level). Obviously, the PHCPs vaccination status was considered when assessing the primary and secondary outcomes of this study.

## BIAS AND LIMITATIONS

The study results will be based on a convenience sample. However the sample will cover a large proportion of geographically well distributed PHCPs.

Selection bias is possibly because of the “late” start of the study: if all the most vulnerable PHCPs have already been infected at the time of the start of this study, then the incidence among the remaining PHCPs may be lower (because better immune system, more adherent to personal protection guidelines etc…). Hence, as in the ongoing seroprevalence study, we will explicitly ask for participation regardless of previous SARS-CoV-2 testing and test results.

Insufficient sample size: due to the current heavy workload in Belgian primary care and time constraints, it might be difficult to recruit PHCPs into this study. However, we will aim for a security margin in the number of participants and have good experience in the ongoing seroprevalence study.

Loss to follow-up or missing data will be possible, for example if a PHCP becomes sick in between two data collection points without providing immediate samples and is isolated at home, or if participant does not provide data at one point because of heavy workload etc. In these cases, the PHCP will be invited to come back in the study and participate in the following data collection time-point. However, in the current outbreak situation PHCPs are supposedly highly interested in knowing their infection status and therefore in participating in the study. Furthermore, their profession might make them more inclined to contribute to medical research. Finally, the duration of follow-up being relatively short, drop out should be minimized. All efforts will be made to maintain the motivation of participants to participate at each time-point by: keeping them regularly updated of the results of the study, being attentive to questions and concerns: keeping communication to a minimum (to avoid overburdening them) and wherever possible communication with participants in their own language.^35^

Under- and overestimation of the presence of SARS-CoV-2 among this population due to imperfect testing methods (imperfect sensitivity and specificity). However, this bias will be minimized by using best available POCT.^19^

### PATIENT & PUBLIC INVOLVEMENT

Patients or the public were not involved in the design, or conduct, or reporting, or dissemination plans of our research. The research team however involved potential study subjects, i.e. PHCPs.

## Supporting information

consent and baseline questionnaire

follow-up questionnaire

## Data Availability

Data are available upon reasonable request. Requests can be directed at both.

https://datastudio.google.com/embed/reporting/7e11980c-3350-4ee3-8291-3065cc4e90c2/page/ZwmOB.

## Ethics and dissemination

Ethical approval has been granted by Ethics Committee of the University Hospital Antwerp/University of Antwerp (Belgian registration number: 3002020000237). Anonymous study results will be made accessible and available as soon as possible after each testing point and at the end of the study to public health authorities involved in management of the COVID-19 epidemic in Belgium. This will be done through a policy brief or press release.

Sciensano will coordinate the distribution of results. These results will also be published on a dedicated, public webpage of the Sciensano COVID-19 dashboard.^36^

The general population will also be informed of the results through press communications. This will be done by the communication departments of the University of Antwerp and the University of Liège, Sciensano and the other study partners.

Scientific peer-reviewed publications (possible short communication, regular paper) will be prepared to add to the body of evidence and availability for the global scientific community and public health decision makers.

## Authors’ contributions

The study concept and design was conceived by SC, NA, BS and ED. SC, NA and BS will conduct registration and data collection. Analysis will be performed by RB. NA prepared the first draft of the manuscript. All authors provided edits and critiqued the manuscript for intellectual content.

## Supplementary materials

Prevalence and incidence of antibodies against SARS-CoV-2 among primary healthcare providers in Belgium –baseline questionnaire

uploaded separately

Prevalence and incidence of antibodies against SARS-CoV-2 among primary healthcare providers in Belgium – Follow-up questionnaire

uploaded separately

