## Supplementary material for "Prevalence and Incidence of Antibodies Against Sars-Cov-2 Among Primary Healthcare Providers in Belgium During One Year of the Covid-19 Epidemic: Prospective Cohort Study Protocol": consent and baseline questionnaire

Dear Participant,

Thank you for your registration for the CHARMING study. We have provided you with your personal study materials for the first three testing time points.

Here we first ask for your formal consent to the study. All questions in the consent section need to be answered before you can proceed. Next we ask for your results on the rapid test, and questions about your health, household, practice and views on the SARS-COV2 pandemic.

Many thanks in advance for carefully completing this questionnaire. We hope this will go smoothly for you.

The CHARMING study team

There are 74 questions in this survey.

### E - Consent

Before giving your consent it is important that you have reviewed the information document about this study available in French **here (https://dox.uliege.be/index.php/s/n64T153cp07BOkG)** and in Dutch **here (https://dox.uliege.be/index.php/s/OYp4cIIx8oxERBt).**

1 Your study code (C followed by 4 numbers; see our email of 25.01.2021 with your link to this questionnaire): *

Please write your answer here:

2
I have received an information sheet (version 2.2, 26-11-2020). All my questions concerning this study have been answered satisfactorily. I was given sufficient time to reflect before agreeing to participate in this study. *

Please choose **only one** of the following:

- Yes

3
My participation is voluntary. I have the right to withdraw my consent at any time without giving a reason. *

Please choose **only one** of the following:

- Yes

4
In order to meet the needs of this study, I consent to the collection and use of my data (including the result of the rapid test). *

Please choose **only one** of the following:

- Yes

## 5
#### I authorise the consultation of my data to the persons collaborating in this research (these persons are listed in the information form). *

Please choose **only one** of the following:

◯ Yes

## 6
#### I agree that the data recorded in this study will be kept for 20 years and may be processed for future research on respiratory infections and coronaviruses. *

Please choose **only one** of the following:

◯ Yes

## 7
#### I agree to provide a blood sample to validate the rapid test. *

Please choose **only one** of the following:

◯ Yes

◯ No

## 8
#### I agree to provide a blood sample to examine the T-cell response. *

Please choose **only one** of the following:

◯ Yes

◯ No

9
I agree that the blood samples taken in this study will be stored for 20 years and can be processed at a later date. *

Please choose **only one** of the following:

- Yes
- No

10
After this study, I agree to be approached for further research. *

Please choose **only one** of the following:

- Yes
- No

11
I wish to participate in this survey. *

Please choose **only one** of the following:

- Yes

### Results of the rapid test

12 *Date on which you carried out the rapid test (dd.mm.yyyy)?* *

Please enter a date:

## 13
#### Did the control line "C" change from blue to red?
#### If not, the test is invalid.

*

Please choose **only one** of the following:

- ◯ Yes
- ◯ No

## 14
#### Result of your quick test for IgG?

#### A red line visible next to G = positive (see figure).
*

Only answer this question if the following conditions are met:
Answer was 'Yes' at question '13 [Q00013]' (Did the control line "C" change from blue to red? If not, the test is invalid. )

Please choose **only one** of the following:

- ◯ Positive
- ◯ Negative
- ◯ Unclear

## 15
#### Result of your quick test for IgM?

#### A red line visible next to M = positive (see figure).

*

Only answer this question if the following conditions are met:
Answer was 'Yes' at question '13 [Q00013]' (Did the control line "C" change from blue to red? If not, the test is invalid. )

Please choose **only one** of the following:

- ◯ Positive
- ◯ Negative
- ◯ Unclear

16 Date on which you completed this questionnaire (dd.mm.yyyy)? *

Please enter a date:

_______________

#### 17 How many sealed tests do you have left after this testing time point? *

❗ Choose one of the following answers
Please choose **only one** of the following:

- ◯ 0 sealed tests
- ◯ 1 sealed test
- ◯ 2 sealed tests
- ◯ 3 sealed tests
- ◯ 4 sealed tests
- ◯ 5 sealed tests

### Your health

## 18

Do you smoke? *

❗ If you choose 'not for________ years' please also specify your choice in the accompanying text field.
❗ Only numbers may be entered in 'not for________ years' accompanying text field.
Please choose **only one** of the following:

- ◯ Yes
- ◯ I have stopped smoking
- ◯ I have never smoked

#### 19 How many years ago did you stop smoking?

Only answer this question if the following conditions are met:
Answer was 'I have stopped smoking' at question '18 [Q00018]' (Do you smoke? )

Please write your answer here:

[ ]

years

## 20

How many alcoholic drinks do you consume per week? *

Please choose **only one** of the following:

- ◯ 0
- ◯ 1 - 5
- ◯ 6 - 10
- ◯ 11 - 15
- ◯ 16 - 20
- ◯ > 20

#### 21 Have you been vaccinated against pneumococcus? *

Please choose **only one** of the following:

- ◯ Yes
- ◯ No
- ◯ I don't know

#### 22 Have you been vaccinated against influenza for this winter season (2020-2021)? *

Please choose **only one** of the following:

- ◯ Yes
- ◯ No
- ◯ I don't know yet

#### 23 Have you been vaccinated against COVID-19? *

Please choose **only one** of the following:

- ◯ Yes
- ◯ No

#### 24 Which vaccine did you receive? *

Only answer this question if the following conditions are met:
Answer was 'Yes' at question '23 [Q00023]' (Have you been vaccinated against COVID-19?)

Please choose **only one** of the following:

- ◯ Pfizer/BioNTech
- ◯ Moderna
- ◯ Oxford/AstraZeneca
- ◯ Other [ ]

#### 25 How many doses have you received? *

Only answer this question if the following conditions are met:
Answer was 'Yes' at question '23 [Q00023]' (Have you been vaccinated against COVID-19?)

Please choose **only one** of the following:

- ◯ 1 dose
- ◯ 2 doses

#### 26 When did you receive the first dose of the vaccine (dd.mm.yyyy)?

Only answer this question if the following conditions are met:
Answer was 'Yes' at question '23 [Q00023]' (Have you been vaccinated against COVID-19?)

Please enter a date:

[ ]

## 27

Do you have one or more chronic diseases? *

Please choose **only one** of the following:

- ◯ Yes
- ◯ No

#### 28 What chronic disease(s) do you have? (multiple answers possible) *

Only answer this question if the following conditions are met:
Answer was 'Yes' at question '27 [Q00027]' (Do you have one or more chronic diseases? )

Please choose **all** that apply:

- ☐ Hypertension
- ☐ Diabetes
- ☐ Obesity
- ☐ Other

#### 29 Please list other chronic diseases

Only answer this question if the following conditions are met:
Answer was at question '28 [Q00028]' (What chronic disease(s) do you have? (multiple answers possible))

Please write your answer here:

#### 30 Do you take medicines for chronic diseases? *

Only answer this question if the following conditions are met:
Answer was 'Yes' at question '27 [Q00027]' (Do you have one or more chronic diseases? )

Please choose **only one** of the following:

- ◯ Yes
- ◯ No

#### 31 If yes which ones? *

Only answer this question if the following conditions are met:
Answer was 'Yes' at question '30 [Q00030]' (Do you take medicines for chronic diseases?)

Please choose **all** that apply:

- ☐ ACE inhibitors
- ☐ Immunosuppressants
- ☐ Corticosteroids (also inhalation)
- ☐ NSAID
- ☐ Other

#### 32 Other medicines for chronic disease

Only answer this question if the following conditions are met:
Answer was at question '31 [Q00031]' (If yes which ones? )

Please write your answer here:

#### 33 Other medicines in the last six months

Only answer this question if the following conditions are met:
Answer was 'Yes' at question ' [Q00034]' (Have you taken medicines other than those for chronic diseases in the last six months?)

Please write your answer here:

#### 34 Have you taken medicines other than those for chronic diseases in the last six months? *

Please choose **only one** of the following:

- ◯ Yes
- ◯ No

### Your general practice

#### 35 I work in general practice as... *

Please choose **only one** of the following:

- ◯ General practitioner
- ◯ General practitioner in training
- ◯ Other healthcare providers, e.g. nurse, dietician, ...

#### 36 Which year of your training are you in?

Only answer this question if the following conditions are met:
Answer was 'General practitioner in training' at question '35 [Q00035]' (I work in general practice as...)

Please choose **only one** of the following:

- Year 1
- Year 2
- Year 3

#### 37 Please select your profession *

Only answer this question if the following conditions are met:
Answer was 'Other healthcare providers, e.g. nurse, dietician, ...' at question '35 [Q00035]' (I work in general practice as...)

Please choose **only one** of the following:

- Nurse
- Psychologist
- Dietician
- Speech therapist
- Other ________

#### 38 I have been doing this job for… *

Please choose **only one** of the following:

- Less than 2 years
- 2 to 5 years
- 6 to 10 years
- More than 10 years

#### 39 I also work at... *

Please choose **all** that apply:

- [ ] As CRA (coordinating and advising doctor)
- [ ] In a hospital
- [ ] In an institution (e.g. psychiatry, care for the disabled, ...)
- [ ] I don't have any other activity
- [ ] Other: ____________

#### 40 Which other healthcare professionals work in your practice? (multiple answers possible) *

Please choose **all** that apply:

- [ ] General practitioner
- [ ] Dietician
- [ ] Psychologist
- [ ] Nurse
- [ ] Practice assistant
- [ ] None of the above
- [ ] Other: ____________

#### 41 What is the (estimated) number of patients assigned to your practice? *

Please write your answer here:

____________

#### 42 What is the (estimated) proportion of patients younger than 15 years of age (%) ? *

❗ Your answer must be between 0 and 100

Please write your answer here:

%

#### 43 What is the (estimated) proportion of patients over 65 years of age (%)? *

❗ Your answer must be between 0 and 100

Please write your answer here:

%

#### 44 What is the estimated proportion of patients with increased benefits (%) ? *

❗ Your answer must be between 0 and 100

Please write your answer here:

%

#### 45 What is the (estimated) proportion of patients with a migration background (%) ? *

Please write your answer here:

%

#### 46 What is the (estimated) proportion of patients who do not speak Dutch, French or German (%) ? *

Please write your answer here:

%

### Your household

#### 47 What is the composition of your household? *

Each answer must be at least 0

Please write your answer(s) here:

How many family members does your household include, including yourself?

How many children attend a crèche (less than 2.5 years) ?

How many children attend pre-school (2,5 to 6 years)?

How many children attend primary school (typically 6 to 12 years)?

How many children attend secondary school (typically 12 - 18 years?

How many household members are university/college students (typically aged over 18 years) AND sleeping in the family home more than 3 nights per week?

How many household members (typically over 18 years) in employment AND sleeping in the family home more than 3 nights per week?

#### 48 Is your partner employed in healthcare with patient contact? *

Please choose **only one** of the following:

- ◯ Yes
- ◯ No
- ◯ Not applicable

#### 49 How many household members had complaints this year that are compatible with COVID-19, including yourself? *

Please write your answer here:

#### 50 If you had complaints, what were they? (multiple answers possible) *

Only answer this question if the following conditions are met:
Answer was greater than or equal to '1' at question '49 [Q00049]' (How many household members had complaints this year that are compatible with COVID-19, including yourself?)

Please choose **all** that apply:

- [ ] I didn't have any complaints
- [ ] Cough
- [ ] Headache
- [ ] Sore throat
- [ ] Fever
- [ ] Shortness of breath
- [ ] Runny nose
- [ ] Muscle pain
- [ ] Loss of sense of smell
- [ ] Loss of taste
- [ ] General weakness/ fatigue
- [ ] Nausea/ vomiting
- [ ] Diarrhoea
- [ ] Other: ____________

#### 51 How many members of your household, including yourself, have been tested for COVID-19 (excluding tests for research purposes)? *

Please write your answer here:

____________

#### 52 How often have you been tested (except for the research purposes)? *

Please write your answer here:

times

#### 53 How many days have you spent in quarantine? *

Please choose **only one** of the following:

- 0 days
- up to 5 days
- up to 7 days
- up to 10 days
- up to 14 days
- up to 20 days
- more than 20 days

#### 54 Have you ever tested positive for COVID-19? *

Please choose **only one** of the following:

- Yes
- No

#### 55 If you tested positive, when was the positive sample taken? *

Only answer this question if the following conditions are met:
Answer was 'Yes' at question '54 [Q00054]' (Have you ever tested positive for COVID-19?)

Please choose **only one** of the following:

- February
- March
- April
- May
- June
- July
- August
- September
- October
- November
- December
- January 2021

#### 56 if you know the exact date of the positive sample enter it here:

Only answer this question if the following conditions are met:
Answer was 'Yes' at question '54 [Q00054]' (Have you ever tested positive for COVID-19?)

Please enter a date:

## 57
#### For the positive test result which test(s) was/were used? (multiple answers possible) *

Only answer this question if the following conditions are met:
Answer was 'Yes' at question '54 [Q00054]' (Have you ever tested positive for COVID-19?)

Please choose **all** that apply:

- [ ] PCR (for virus detection)
- [ ] Rapid test (for virus detection)
- [ ] Blood sample (for antibody detection)
- [ ] Rapid test (for antibody detection)
- [ ] Other: ______

## 58
#### If you tested positive, who was the suspected source of the infection? *

Only answer this question if the following conditions are met:
Answer was 'Yes' at question '54 [Q00054]' (Have you ever tested positive for COVID-19?)

Please choose **all** that apply:

- [ ] Patient
- [ ] Co-worker
- [ ] Family member
- [ ] Other: ______

#### 59 If you were treated for COVID-19, what treatment did you have? *

Only answer this question if the following conditions are met:
Answer was 'Yes' at question '54 [Q00054]' (Have you ever tested positive for COVID-19?)

Please choose **all** that apply:

- ☐ Symptomatic treatment of pain, fever and other complaints
- ☐ Hydroxychloroquine
- ☐ Antibiotics
- ☐ No treatment
- ☐ Other: ____________

#### 60 If you were admitted for COVID-19, how many days did you spend in hospital?

#### (if you were not admitted to hospital put '0') *

Only answer this question if the following conditions are met:
Answer was 'Yes' at question '54 [Q00054]' (Have you ever tested positive for COVID-19?)

Please write your answer here:

____________

days

## 61
#### If you were admitted for COVID-19, how many days did you stay in intensive care? (if you were not admitted to intensive care put '0') *

Only answer this question if the following conditions are met:
Answer was 'Yes' at question '54 [Q00054]' (Have you ever tested positive for COVID-19?)

Please write your answer here:

days

#### 62 How many household members have tested positive for COVID-19, **not** including yourself? *

Please write your answer here:

#### 63 How many household members have been admitted to hospital for (suspected) COVID-19, **not** including yourself? *

Please write your answer here:

#### 64 How many household members have been treated for (suspected) COVID-19, **not** including yourself? *

Please write your answer here:

### Risk factors for COVID-19

#### 65 Have you continued to work since the outbreak? *

Please choose **only one** of the following:

- Yes
- No

#### 66 Have you been in physical contact with patients with confirmed COVID-19 since the outbreak? *

Please choose **only one** of the following:

- Yes
- No

#### 67 If so, how many? *

Only answer this question if the following conditions are met:
Answer was 'Yes' at question '66 [Q00066]' (Have you been in physical contact with patients with confirmed COVID-19 since the outbreak?)

Please choose **only one** of the following:

- 1 - 5 patients
- 6 - 10 patients
- 11 - 15 patients
- 16 - 20 patients
- > 20 patients

#### 68 Have you lacked protective equipment since the outbreak? *

Please choose **only one** of the following:

- Yes
- No

#### 69 If so which equipment? (multiple answers possible) *

Only answer this question if the following conditions are met:
Answer was 'Yes' at question '68 [Q00068]' (Have you lacked protective equipment since the outbreak?)

Please choose **all** that apply:

- Gloves
- Surgical mouth mask
- Other mouth mask (FFP2 or FFP3)
- Safety goggles
- Apron / body protection
- Other:

#### 70 If available, which protective material do you use in patients with (suspected) COVID-19)? (multiple answers possible) *

Please choose **all** that apply:

- Gloves
- Surgical mouth mask
- Other mouth mask (FFP2 or FFP3)
- Safety goggles
- Apron/body protection
- Other:

#### 71 If available, what protective material do you use with your other patients? (multiple answers possible) *

Please choose **all** that apply:

- ☐ Gloves
- ☐ Surgical mouth mask
- ☐ Other mouth mask (FFP2 or FFP3)
- ☐ safety goggles
- ☐ Apron/body protection
- ☐ Other: ________

#### 72 Have you participated in the COVID patient triage? *

Please choose **only one** of the following:

- ◯ Yes
- ◯ No

#### 73 If so, how many patients did you physically examine who subsequently turned out to be COVID-19 positive? *

Only answer this question if the following conditions are met:
Answer was 'Yes' at question '72 [Q00072]' (Have you participated in the COVID patient triage?)

Please choose **only one** of the following:

- ◯ 0 patients
- ◯ 1 - 5 patients
- ◯ 6 - 10 patients
- ◯ 11 - 15 patients
- ◯ 16 - 20 patients
- ◯ > 20 patients

## 74
#### Indicate to what extent you agree with the following statements

#### (1= totally disagree; 5= totally agree): *

Please choose the appropriate response for each item:

| | 1 | 2 | 3 | 4 | 5 |
|---|---|---|---|---|---|
| **The personal protection equipment that I use, protects me sufficiently against more contagious variants of SARS-CoV-2.** | ○ | ○ | ○ | ○ | ○ |
| **A temporary ban on non-essential international travel is needed.** | ○ | ○ | ○ | ○ | ○ |
| **I am sure I am already infected with COVID-19.** | ○ | ○ | ○ | ○ | ○ |
| **I will certainly be infected with COVID-19 during this epidemic.** | ○ | ○ | ○ | ○ | ○ |
| **I am afraid I am contaminating my relatives.** | ○ | ○ | ○ | ○ | ○ |
| **The guidelines for primary care are clearly communicated.** | ○ | ○ | ○ | ○ | ○ |
| **The guidelines for primary care are scientifically based.** | ○ | ○ | ○ | ○ | ○ |
| **The Belgian healthcare system is strong enough to cope with this epidemic.** | ○ | ○ | ○ | ○ | ○ |
| **The testing capacity in Belgium is sufficient.** | ○ | ○ | ○ | ○ | ○ |
| **Rapid diagnostic tests are relevant for general practice.** | ○ | ○ | ○ | ○ | ○ |
| **Rapid diagnostic tests for SARS-CoV-2 viral detection are manageable for general practice.** | ○ | ○ | ○ | ○ | ○ |

| | 1 | 2 | 3 | 4 | 5 |
|---|---|---|---|---|---|
| **The measures imposed by the government are sufficient.** | ◯ | ◯ | ◯ | ◯ | ◯ |
| **Everyone should wear a mask if they go outdoors.** | ◯ | ◯ | ◯ | ◯ | ◯ |
| **I have every confidence in the scientific COVID-19 expert committee.** | ◯ | ◯ | ◯ | ◯ | ◯ |
| **Most of my patients follow the rules of 'social distancing'.** | ◯ | ◯ | ◯ | ◯ | ◯ |
| **Most of my patients adhere to hygiene rules.** | ◯ | ◯ | ◯ | ◯ | ◯ |
| **La plupart de mes patients symptomatiques respectent les règles de quarantaine.** | ◯ | ◯ | ◯ | ◯ | ◯ |
| **This period is more stressful than during a busy flu period.** | ◯ | ◯ | ◯ | ◯ | ◯ |
| **I want to get the COVID-19 vaccination as soon as it is available.** | ◯ | ◯ | ◯ | ◯ | ◯ |

Thank you very much for completing this questionnaire.

You will shortly receive an email that will explain what your test result means. We will send you an overview of your consent to participate in the study in the coming weeks.

The CHARMING study team

CHARMING

Coronavirus HuisARtsenpraktijk - MédecINe Générale

Submit your survey.
Thank you for completing this survey.
