## Supplementary material for "Prevalence and Incidence of Antibodies Against Sars-Cov-2 Among Primary Healthcare Providers in Belgium During One Year of the Covid-19 Epidemic: Prospective Cohort Study Protocol": follow-up questionnaire

Dear Participant,

Thank you for your participation in CHARMING. This follow-up questionnaire refers to the period since the last testing period.

Many thanks in advance for carefully completing this questionnaire. We hope this will go smoothly for you.

The CHARMING study team

CHARMING
Coronavirus HuisARtsenpraktijk - MédecINe Générale

There are 46 questions in this survey.

#### Part 1

1 Your personal study code (C followed by 4 numbers; see our email of 26.02.2021 with your link to this questionnaire): *

Please write your answer here:

#### Part 2

Instructions on how to perform the rapid test can be found in French **here** (https://dox.uliege.be/index.php/s/1duglah08HN8YIr) and in Dutch **here** (https://dox.uliege.be/index.php/s/hqqiswSGBxKw3yf). Short instruction videos are available here:

- French test on yourself : https://vimeo.com/492411023/7b2bedb700 (https://vimeo.com/492411023/7b2bedb700)
- French test on someone else: https://vimeo.com/492427669/b42bb624b6 (https://vimeo.com/492427669/b42bb624b6)
- Dutch test on yourself: https://vimeo.com/492430777/92626224d1 (https://vimeo.com/492430777/92626224d1)
- Dutch test on someone else : https://vimeo.com/492428827/d565f20bc2 (https://vimeo.com/492428827/d565f20bc2)

2 *Date on which you carried out the rapid test (dd.mm.yyyy)?* *

Please enter a date:

3

Did the control line "C" change from blue to red?

If not, the test is invalid.

*

Please choose **only one** of the following:

◯ Yes

◯ No

4
Result of your quick test for IgG?

A red line visible next to G = positive (see figure).
*

Please choose **only one** of the following:

- ◯ Positive
- ◯ Negative
- ◯ Unclear

5
Result of your quick test for IgM?

A red line visible next to M = positive (see figure).
*

Please choose **only one** of the following:

- ◯ Positive
- ◯ Negative
- ◯ Unclear

6 Date on which you completed this questionnaire (dd.mm.yyyy)? *

Please enter a date:

##### 7 How many sealed tests do you have left after this testing time point? *

Please choose **only one** of the following:

- ◯ 0 sealed tests
- ◯ 1 sealed test
- ◯ 2 sealed tests
- ◯ 3 sealed tests
- ◯ 4 sealed tests
- ◯ 5 sealed tests

### Part 3

##### 8 Since **your first** testing period (end December 2020 or end January 2021), how many days have you spent in quarantine? *

Please choose **only one** of the following:

- ◯ 0 days
- ◯ up to 5 days
- ◯ up to 7 days
- ◯ up to 10 days
- ◯ up to 14 days
- ◯ up to 20 days
- ◯ more than 20 days

#### 9 Since **your first** testing period (end December 2020 or end January 2021), how often have you been tested for COVID-19 (except for research purposes)? *

Please write your answer here:

times

#### 10 I work in general practice as... *

Please choose **only one** of the following:

- General practitioner
- General practitioner in training
- Other healthcare providers, e.g. nurse, dietician, ...

#### 11 Please select your profession *

Please choose **only one** of the following:

- Nurse
- Psychologist
- Dietician
- Speech therapist
- Other

#### 12 Since the **last** testing period I have also worked... *

Please choose **all** that apply:

- ☐ As CRA (coordinating and advising doctor)
- ☐ In a hospital
- ☐ In an institution (e.g. psychiatry, care for the disabled, ...)
- ☐ I don't have any other activity
- ☐ Other: __________

### Part 4

#### 13 Since the last testing phase of CHARMING how many family members had complaints that are compatible with COVID-19, including yourself? *

Please write your answer here:

__________

#### 14 If you had complaints, since the last testing period, what were they? (multiple answers possible) *

Please choose **all** that apply:

- [ ] I didn't have any complaints
- [ ] Cough
- [ ] Headache
- [ ] Sore throat
- [ ] Fever
- [ ] Shortness of breath
- [ ] Runny nose
- [ ] Muscle pain
- [ ] Loss of sense of smell
- [ ] Loss of taste
- [ ] General weakness/ fatigue
- [ ] Nausea/ vomiting
- [ ] Diarrhoea
- [ ] Other: ______

#### 15 Since the last testing period how many family members, including yourself, have been tested for COVID-19 (excluding tests for research purposes)? *

Please write your answer here:

______

#### 16 Have you tested positive for COVID-19 since the last testing period? (multiple answers possible) *

Please choose **only one** of the following:

- ◯ Yes
- ◯ No

## 17
#### For the positive test result which test(s) was/were used? (multiple answers possible) *

Please choose **all** that apply:

- ☐ PCR (for virus detection)
- ☐ Rapid test (for virus detection)
- ☐ Blood sample (for antibody detection)
- ☐ Rapid test (for antibody detection)
- ☐ Other: __________

#### 18 If you tested positive when was the positive sample taken (dd.mm.yyyy)?

Please enter a date:

__________

#### 19 If you were treated for COVID-19, what treatment did you have? *

Please choose **all** that apply:

- ☐ Symptomatic treatment of pain, fever and other complaints
- ☐ Hydroxychloroquine
- ☐ Antibiotics
- ☐ No treatment
- ☐ Other: ______

## 20
#### If you tested positive, who was the suspected source of the infection? *

Please choose **all** that apply:

- ☐ Patient
- ☐ Co-worker
- ☐ Family member
- ☐ Other: ______

## 21
#### If you were admitted for COVID-19, how many days did you spend in hospital?

#### (if you were not admitted to hospital put '0') *

Please write your answer here:

______

days

22
If you were admitted for COVID-19, how many days did you stay in intensive care? (if you were not admitted to intensive care put '0') *

Please write your answer here:

days

23 Since the last testing period how many family members have tested positive for COVID-19, **not** including yourself? *

Please write your answer here:

24 Since the last testing period how many family members have been admitted to hospital for (suspected) COVID-19, **not** including yourself? *

Please write your answer here:

25 Since the last testing period how many family members have been treated for (suspected) COVID-19, **not** including yourself? *

Please write your answer here:

### Part 5

#### 26 Have you continued to work in primary care since the **last** testing period? *

Please choose **only one** of the following:

- Yes
- No

#### 27 Have you been in physical contact with patients with confirmed COVID-19 since the last testing period? *

Please choose **only one** of the following:

- Yes
- No

#### 28 If so, how many? *

Please choose **only one** of the following:

- 1 - 5 patients
- 6 - 10 patients
- 11 - 15 patients
- 16 - 20 patients
- > 20 patients

#### 29 Have you lacked protective equipment since the **last** testing period? *

Please choose **only one** of the following:

- ◯ Yes
- ◯ No

#### 30 If so which equipment? (multiple answers possible) *

Please choose **all** that apply:

- ☐ Gloves
- ☐ Surgical mouth mask
- ☐ Other mouth mask (FFP2 or FFP3)
- ☐ Safety goggles
- ☐ Apron / body protection
- ☐ Other: ____________

#### 31 If available, which protective material have you used since the last testing period in patients with (suspected) COVID-19)? (multiple answers possible) *

Please choose **all** that apply:

- ☐ Gloves
- ☐ Surgical mouth mask
- ☐ Other mouth mask (FFP2 or FFP3)
- ☐ Safety goggles
- ☐ Apron/body protection
- ☐ Other: ____________

## 32
#### If available, what protective material have you used with your other patients? (multiple answers possible) *

Please choose **all** that apply:

- ☐ Gloves
- ☐ Surgical mouth mask
- ☐ Other mouth mask (FFP2 or FFP3)
- ☐ safety goggles
- ☐ Apron/body protection
- ☐ Other: ____________

#### 33 Have you participated in the COVID patient triage since the last testing period? *

Please choose **only one** of the following:

- ○ Yes
- ○ No

#### 34 If so, how many patients did you physically examine who subsequently turned out to be COVID-19 positive? *

Please choose **only one** of the following:

- ○ 0 patients
- ○ 1 - 5 patients
- ○ 6 - 10 patients
- ○ 11 - 15 patients
- ○ 16 - 20 patients
- ○ > 20 patients

#### 35 Have you been vaccinated against COVID-19? *

Please choose **only one** of the following:

- ◯ Yes
- ◯ No

#### 36 Which vaccine did you receive? *

Please choose **only one** of the following:

- ◯ Pfizer/BioNTech
- ◯ Moderna
- ◯ Oxford/AstraZeneca
- ◯ Other [ ]

#### 37 How many doses have you received? *

Please choose **only one** of the following:

- ◯ 1 dose
- ◯ 2 doses

#### 38 When did you receive the **first** dose of the vaccine (dd.mm.yyyy)? *

Please enter a date:

[ ]

#### 39 Did you experience side-effects after receiving the **first** dose? *

Please choose **only one** of the following:

- ◯ No side-effects
- ◯ Negligible side-effects
- ◯ Mild side-effects
- ◯ Moderate side-effects
- ◯ Severe side-effects

#### 40 For how many days did you experience the following side-effects after the **first** dose (if you did not experience the side-effect put '0'): *

#### 41 What other moderate or severe side-effects did you experience after receiving the **first** dose?

Please write your answer here:

#### 42 When did you receive the **second** dose of the vaccine (dd.mm.yyyy)? *

Please enter a date:

#### 43 Did you experience side-effects after receiving the **second** dose? *

Please choose **only one** of the following:

- ◯ No side-effects
- ◯ Negligible side-effects
- ◯ Mild side-effects
- ◯ Moderate side-effects
- ◯ Severe side-effects

#### 44 For how many days after receiving the **second** dose of the vaccine did you experience the following side-effects (if you did not experience the side-effect put '0')? *

#### 45 What other moderate or severe side-effects did you experience after receiving the **second** dose?

Please write your answer here:

## 46
#### Indicate to what extent you agree with the following statements

#### (1= totally disagree; 5= totally agree): *

Please choose the appropriate response for each item:

| | 1 | 2 | 3 | 4 | 5 |
|---|---|---|---|---|---|
| **The personal protection equipment that I use, protects me sufficiently against more contagious variants of SARS-CoV-2.** | ○ | ○ | ○ | ○ | ○ |
| **A temporary ban on non-essential international travel is still needed.** | ○ | ○ | ○ | ○ | ○ |
| **The Belgian healthcare system is strong enough to cope with this epidemic.** | ○ | ○ | ○ | ○ | ○ |
| **The testing capacity in Belgium is sufficient.** | ○ | ○ | ○ | ○ | ○ |
| **Rapid diagnostic tests are relevant for general practice.** | ○ | ○ | ○ | ○ | ○ |
| **Rapid diagnostic tests for SARS-CoV-2 viral detection are manageable for general practice.** | ○ | ○ | ○ | ○ | ○ |
| **The measures imposed by the government are sufficient.** | ○ | ○ | ○ | ○ | ○ |
| **Everyone should wear a mask when they work inside with other people.** | ○ | ○ | ○ | ○ | ○ |
| **I have every confidence in the scientific COVID-19 expert committee.** | ○ | ○ | ○ | ○ | ○ |
| **Most of my patients follow the rules of 'social distancing'.** | ○ | ○ | ○ | ○ | ○ |
| **Most of my patients adhere to hygiene rules.** | ○ | ○ | ○ | ○ | ○ |

| | 1 | 2 | 3 | 4 | 5 |
|---|---|---|---|---|---|
| **Most of my symptomatic patients respect the quarantine rules.** | ○ | ○ | ○ | ○ | ○ |
| **This period is more stressful than during a busy flu period.** | ○ | ○ | ○ | ○ | ○ |
| **For health care personnel the COVID-19 vaccination should be obligatory.** | ○ | ○ | ○ | ○ | ○ |

Thank you very much for completing this questionnaire.

You will shortly receive an email that will explain what your test result means.

If you experience side-effects after receiving the vaccination you can report them officially here:

In Dutch: https://www.fagg.be/nl/bijwerking (https://www.fagg.be/nl/bijwerking)
In French: https://www.afmps.be/fr/effet_indesirable (https://www.afmps.be/fr/effet_indesirable)

The CHARMING study team

CHARMING
Coronavirus HuisARtsenpraktijk - MédecINe Générale

21.03.2021 – 20:58

Submit your survey.
Thank you for completing this survey.
